# Automated Eye-Tracking for Parkinson’s Disease Diagnosis: A Proof-of-Concept Cascade Classifier Study Establishing Clinical Validity

**DOI:** 10.64898/2026.06.22.26355826

**Authors:** J. Michael Menke, Sana Aslam, Hector Rieiro, Ron Waldorf, Holly A. Shill

## Abstract

**Background:** Parkinson’s disease (PD) is a progressive neurodegenerative disorder of increasing prevalence, with diagnostic accuracy of approximately 26% in early symptomatic patients. There is a need for accurate, non-invasive biomarkers to aid in disease diagnosis.

**Methods:** This proof-of-concept study enrolled 90 participants (PD n = 30, other movement disorders [OM] n = 30, healthy controls [HC] n = 30) at a single institution. Participants completed two 10-minute eye-tracking sessions using the SaccadeDX™ 250-Hz binocular system. A two-level cascade classifier was fitted using elastic-net feature selection followed by logistic regression on the selected features, validated by 10-fold cross-validation. The cascade distinguished HC from movement disorders (Level 1) and PD from OM (Level 2), with the objective of establishing clinical validity — that an eye-tracking signal correlates reliably with PD diagnosis.

**Results:** Level 1 achieved an area under the curve (AUC) of 0.818 (95% CI: 0.71, 0.91), with a sensitivity of 83% and specificity of 63%. Level 2 achieved an AUC of 0.670 (95% CI: 0.52, 0.80), with a sensitivity of 68% and specificity of 63%. End-to-end PD detection achieved an AUC of 0.866 and an accuracy of 83.5%, meeting the prospectively specified accuracy threshold and the proof-of-concept AUC benchmark. Five adverse events were recorded (three cases of dizziness, one of nausea, and one of dry eyes); one participant withdrew from the study.

**Conclusions:** Clinical validity is established: a reproducible eye-tracking signal for PD is detectable using a two-level cascade classifier. A multi-center confirmatory study is warranted before assessment of clinical utility.

**Trial registration:** ClinicalTrials.gov NCT04925622.

## 1. Introduction

Parkinson’s disease (PD) is a progressive neurodegenerative disease that is increasing in frequency. It represents a substantial impact on quality of life and carries a significant economic burden worldwide [9,26]. The clinical diagnosis of PD can be challenging, with studies suggesting approximately 26% accuracy in those with short duration of symptoms and/or prior to medication treatment [2]. Following clinical progression and response to dopaminergic therapies improves diagnostic accuracy to approximately 90%, but often requires many years of follow-up [2,3,13]. Dopamine transporter imaging (e.g., DaTscan) can improve diagnostic accuracy in patients with early symptoms but does not distinguish between types of degenerative parkinsonism, is costly, invasive, and requires specialized neuroimaging centers.

More recently, detection of α-synuclein in skin and cerebrospinal fluid (CSF) appears promising, even in early disease, but may be limited by access to specialty laboratories and patient reluctance to undergo lumbar puncture [6,22]. There is a need for non-invasive biomarker testing that is easy to administer across large populations of patients. Automated testing of eye movements has recently shown promise in the discrimination of PD either on its own [15] or in combination with other biomarkers [7,20].

A recent review of eye tracking in PD summarized the field around four recurring oculomotor domains: saccadic abnormalities, fixation instability, smooth-pursuit deficits, and pupillary changes. The review emphasized that these measures are objective, non-invasive, and potentially useful not only for diagnosis, but also for disease monitoring, cognitive assessment, and rehabilitation [7]. It also noted, however, that clinical translation depends on standardized protocols, validation of predictive models, and integration of eye tracking into practical clinical workflows [7].

SaccadeDX™ (Saccadous Inc., Scottsdale, AZ) is an eye-tracking system that uses a proprietary algorithm to capture and process microscopic eye movements using a virtual reality headset, providing non-invasive measurement in approximately 20 minutes. Eye movements provide a promising window into neural function in PD. Saccadic hypometria, square-wave jerk (SWJ) elevation, and smooth-pursuit deficits are each linked to defined basal ganglia and brainstem circuit pathology (Figure 1) [4,27]. Movement disorders such as progressive supranuclear palsy (PSP), corticobasal degeneration (CBD), and essential tremor present with characteristic oculomotor profiles, creating a diagnostic landscape that rewards objective, quantitative measurement.

**Figure 1.**
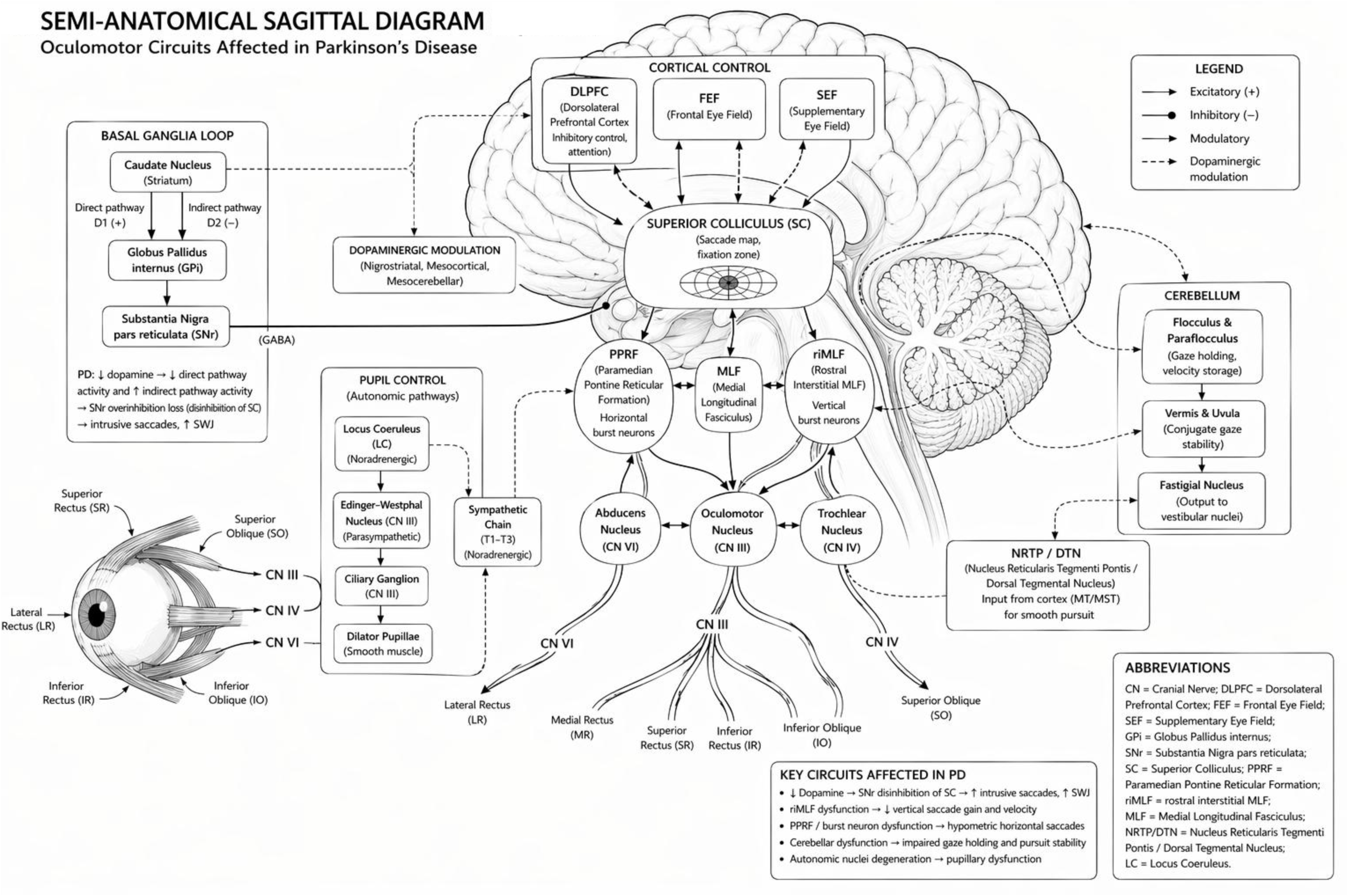
Neuroanatomic basis of eye-movement features. Sagittal view of brainstem and cerebellar circuits underlying the saccade, smooth-pursuit, fixation, and optokinetic measurements analyzed in this study.

We undertook this proof-of-concept study to establish the clinical validity of SaccadeDX™ for PD detection — that is, to demonstrate that a reliable eye-tracking signal for PD exists and can be detected with a reproducible method — using a two-level cascade classifier with elastic-net feature selection and 10-fold cross-validated logistic regression. This approach yields a fully auditable scoring equation and conservative cross-validated performance estimates.

## 2. Methods

### 2.1 Study Design and Participants

Using an IRB-approved protocol (NCT04925622), we enrolled a convenience sample of participants at the Barrow Neurological Institute between January 2021 and April 2022 (IRB # PHX-20-500-177-80-04 approved November 10, 2020). The study was approved by St Joseph’s Hospital and Medical Center Institutional Review Board (Phoenix, AZ) and conducted in accordance with the Declaration of Helsinki. Sample size (n = 30 per group) was selected to detect clinically relevant group differences.

Patients with Parkinson’s disease (PD) met MDS diagnostic criteria for established PD [12,19]. Non-PD movement disorder patients (OM) included those with essential tremor, PSP, dystonia, Huntington’s disease, ataxia, CBD, and tardive dyskinesia. Age- and sex-matched healthy controls (HC) were free of neurological disease. All participants were reviewed by study investigators for clinical diagnosis and appropriateness for inclusion.

Exclusion criteria included significant concurrent drug or alcohol use, unstable medical problems, Hoehn-Yahr stage 4 or higher (parkinsonism participants only), dementia, inability to complete the protocol, and primary vision problems such as macular degeneration.

Testing was performed in the clinic setting, typically after a routine clinic visit. Participants were seated in a chair and the virtual reality goggles were placed on the head. The paradigm was presented twice with a 10-minute rest between sessions; session measurements were averaged. Total testing time was approximately 20 minutes. Adverse events were ascertained throughout the study.

### 2.2 Feature Engineering and Preprocessing

Eye-tracking data from 90 participants yielded 75 raw saccadic measurements across six task domains. Missing values were addressed by multiple imputation by chained equations (MICE, predictive mean matching, 10 iterations) [23]. Features were winsorized at the 1st/99th percentile and z-score standardized. Twenty-two engineered features targeting PD-specific neural pathway signatures were added, giving 97 candidate predictors.

### 2.3 Cascade Classifier

Each cascade level was fitted in two stages. First, elastic-net regularized logistic regression (α = 0.5) selected an informative subset of features by 10-fold cross-validated penalty selection at λ_1se_ using glmnet [8]. Features with non-zero coefficients at λ_1se_(|β| > 1×10^−3^) were retained. Second, a logistic regression model was fitted on the selected features. Level 1 distinguished HC from any movement disorder; Level 2 distinguished PD from OM.

This pipeline was selected a priori as the analytic plan for this manuscript. An earlier exploratory analysis of the same dataset, in which a large grid of model configurations was searched, produced an apparently higher Level-2 AUC and is not the basis for inference here. The cross-validated logistic-regression analysis provides conservative, replication-oriented performance estimates appropriate for a clinical-validity claim.

### 2.4 Validation

Model performance was evaluated using 10-fold cross-validation. At each iteration, the logistic regression model was fitted on the training subset and tested on the held-out subset to generate out-of-fold predicted probabilities. Fold assignment preserved class balance across groups. Receiver-operating characteristic (ROC) curves, area under the curve (AUC), and 95% confidence intervals were estimated from the resulting out-of-fold probabilities using bootstrap resampling (2,000 iterations).

Before analysis, proof-of-concept success criteria were defined as overall classification accuracy ≥ 0.75, consistent with the prospectively registered primary outcome measure in ClinicalTrials.gov (NCT04925622), which specified accurate diagnosis in more than 75% of patients. An end-to-end AUC benchmark of ≥ 0.80 was additionally used as a proof-of-concept performance target representing clinically meaningful discrimination.

### 2.5 Unified Scoring and Thresholds

Each participant received a single cascade score S combining the two classifier levels:

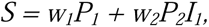

where P_1_ is the Level-1 out-of-fold probability of any movement disorder (healthy control vs. disorder), P_2_ is the Level-2 out-of-fold probability of Parkinson’s disease (PD vs. other movement disorder), and I_1_ is an indicator equal to 1 when P_1_ ≥ 0.50 and 0 otherwise. The weights w_1_ = 0.45 and w_2_ = 0.55 give modestly greater influence to the PD-specific Level-2 probability, but only when Level-1 has first flagged the participant as likely abnormal. Participants who do not clear the Level-1 threshold retain a score of S = w_1_P_1_, which preserves healthy-control separation without forcing a PD probability onto controls.

Two operating cutoffs were applied downstream: S = 0.50 for the Level-1 (HC vs. movement-disorder) decision and S = 0.62 for the Level-2 (PD vs. OM) decision. Calibration curves for group-membership probabilities were estimated by bootstrap-resampled logistic regression (n = 2,000).

The Level-1 threshold (S = 0.50) was selected because it represents equal posterior odds of healthy versus movement-disorder classification. The Level-2 threshold (S = 0.62) corresponded to the midpoint of the observed PD and OM score distributions and approximated the operating point maximizing balanced sensitivity and specificity in cross-validated analyses. These thresholds should be regarded as provisional and subject to refinement in future validation studies.

## 3. Results

### 3.1 Participants

There were 96 participants screened for the study. Three did not meet eligibility criteria, two were unable to complete due to technical issues, and one did not complete due to adverse events. This left 90 participants analyzed: PD n = 30, OM n = 30, HC n = 30.

Within the OM group: essential tremor n = 11, Huntington’s disease n = 7, ataxia n = 4, PSP n = 3, dystonia n = 3, CBD n = 1, and tardive dyskinesia n = 1. Groups were comparable in age (PD 67.1 ± 10.6 yr; OM 60.9 ± 13.0 yr; HC 67.1 ± 10.9 yr) and sex (PD/HC 57% male; OM 47% male).

Adverse events occurred in five participants: three reported dizziness (one discontinued the study), one nausea, and one dry eyes. No serious adverse events were recorded.

### 3.2 Feature Selection and Model Coefficients

Elastic-net selection retained 10 features for Level 1 and 14 features for Level 2 from the 97 candidates. The principal discriminating features are summarized clinically in Supplementary Tables S1 and S2, which list the feature name, expected clinical interpretation, relevant neuroanatomic circuit, and approximate contribution to classification. The full list of candidate features and their selection status is provided in Supplementary Table S3.

### 3.3 Cascade Classifier Performance

Level 1: 10-fold cross-validated AUC = 0.818 (95% CI: 0.71, 0.91), sensitivity 83.4%, specificity 63.3%. At 30% movement-disorder prevalence: PPV 48%, NPV 91%.

Level 2: 10-fold cross-validated AUC = 0.670 (95% CI: 0.52, 0.80), sensitivity 67.7%, specificity 63.3%. At 50% conditional prevalence: PPV 64.9%, NPV 66.3%.

Receiver-operating characteristic curves and PPV/NPV functions for both cascade levels are shown in Figure 2.

**Figure 2.**
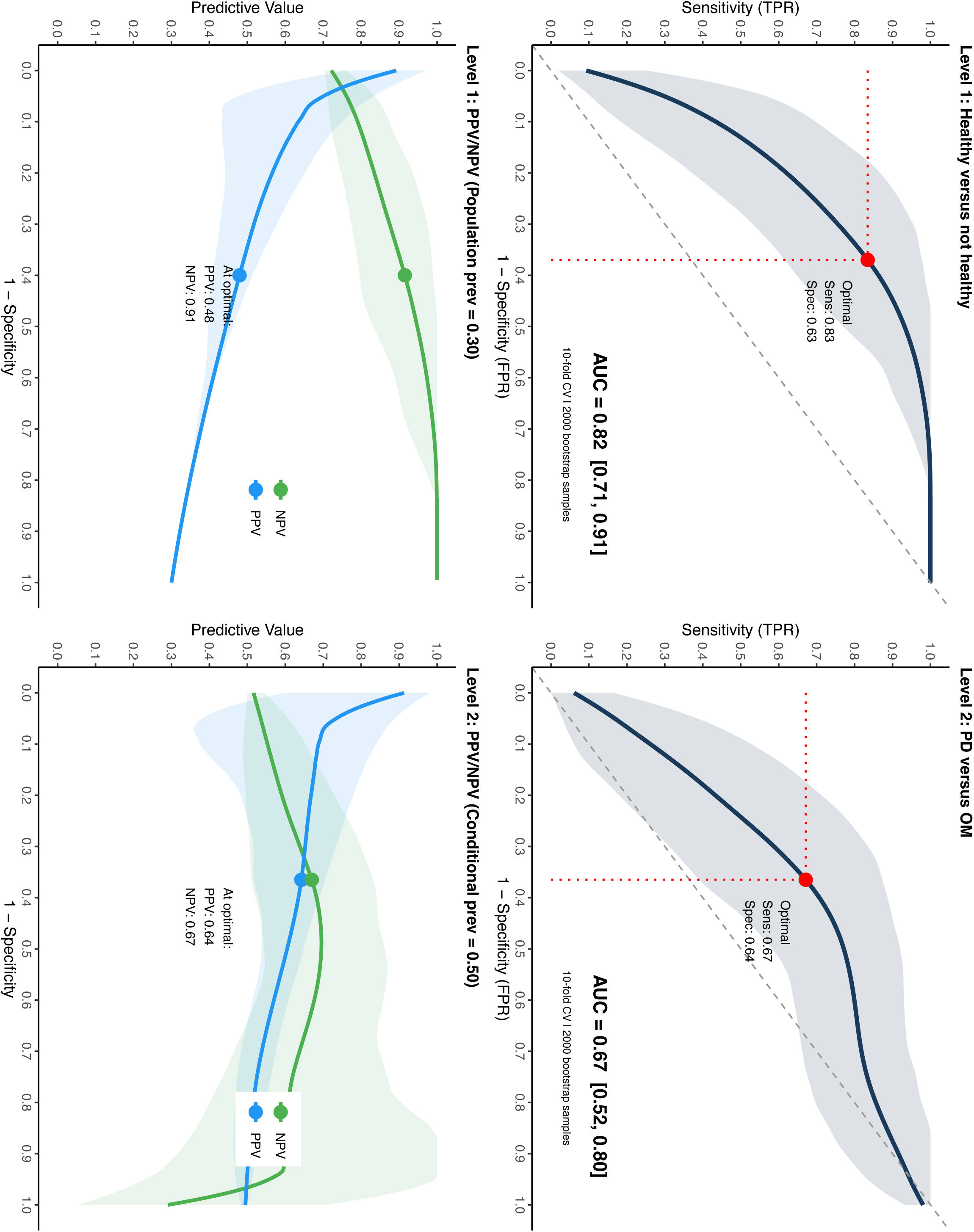
Diagnostic performance of the cascade classifier. ROC curves with 10-fold cross-validation bootstrap 95% confidence bands (left) and predictive-value functions (right) for Level 1 (HC vs. any movement disorder, top row) and Level 2 (PD vs. OM, bottom row). Level 1 AUC = 0.818 (95% CI: 0.71, 0.91); Level 2 AUC = 0.670 (95% CI: 0.52, 0.80). PPV/NPV are plotted across plausible prevalence ranges for each cascade level.

### 3.4 End-to-End PD Detection

End-to-end AUC = 0.866. Cascade hard classification achieved accuracy 83.5%, sensitivity 90.3%, specificity 98.3%, PPV 96.6%, and NPV 95.2%. The prospectively specified accuracy threshold (≥ 0.75) and the proof-of-concept AUC benchmark (≥ 0.80) were both met.

Because the unified cascade score incorporates information from both classifier levels and preserves healthy-control separation through the Level-1 gate, end-to-end discrimination is not expected to equal the performance of either classifier level considered in isolation.

Receiver-operating characteristic curves and predictive-value functions for both cascade levels are shown in Figure 2. Density distributions of the unified cascade score across HC, OM, and PD groups are shown in Figure 3, and calibration curves relating the unified score to estimated group-membership probabilities are shown in Figure 4.

**Figure 3.**
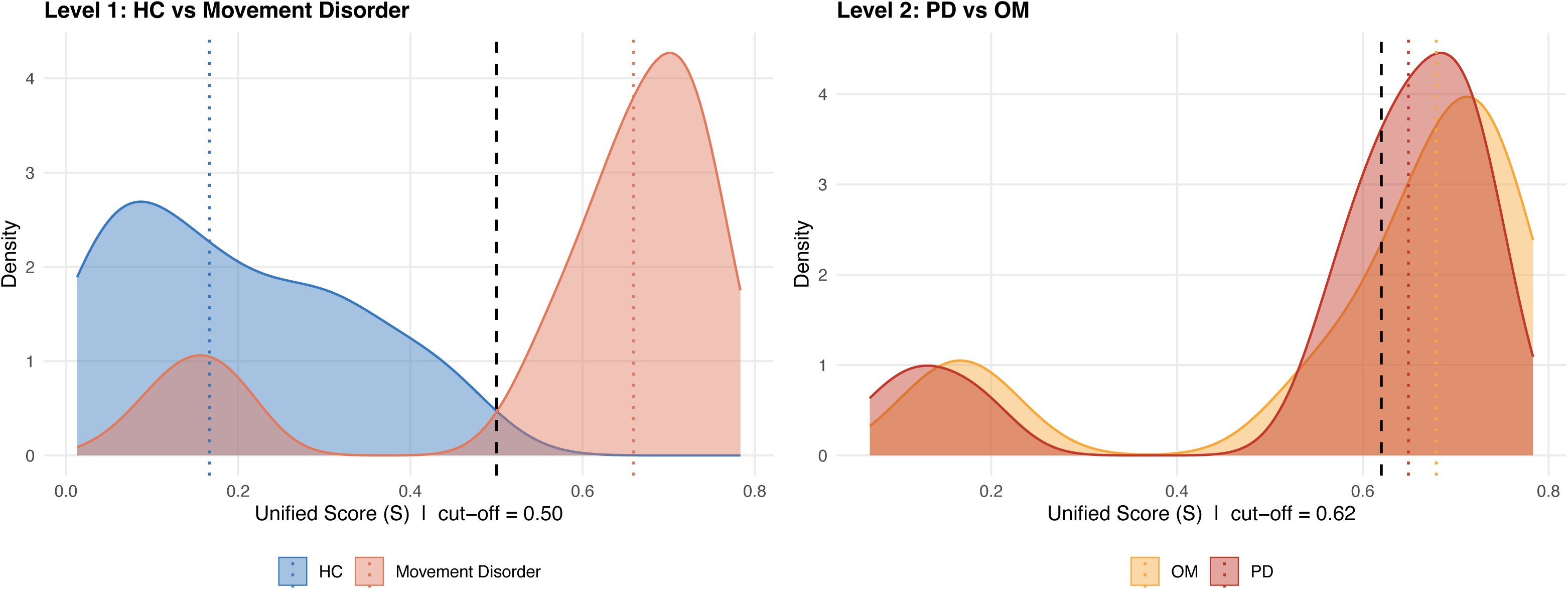
Density of the unified score S by clinical group. Left: Level 1 (HC vs. movement disorder, cut-off S = 0.50). Right: Level 2 (PD vs. OM, cut-off S = 0.62).

**Figure 4.**
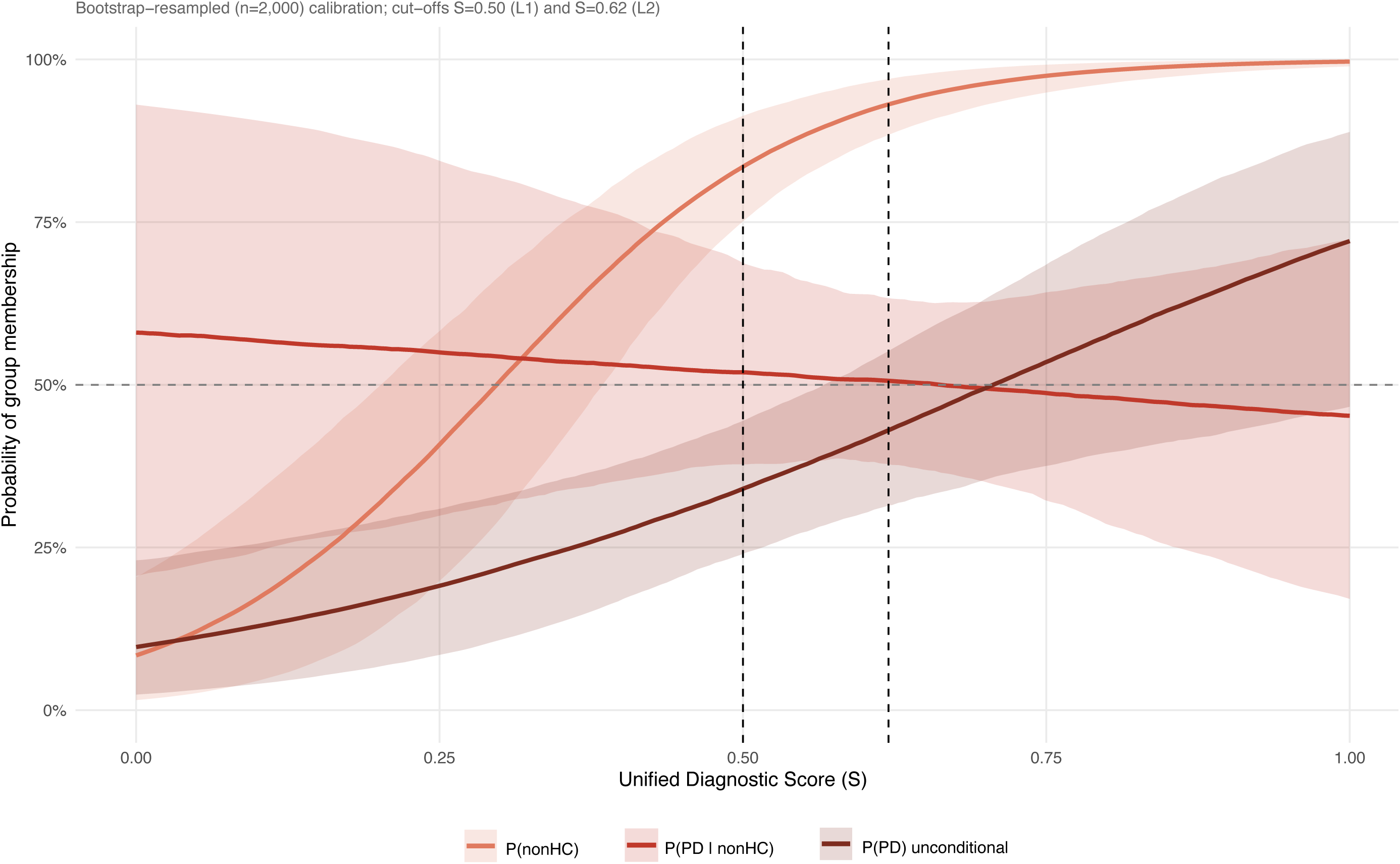
Exploratory group membership probability calibration.

### 3.5 Clinical Interpretation of the Scoring Algorithm

Lower composite scores were strongly associated with healthy-control classification, whereas higher scores were associated with Parkinson’s disease classification. Scores below 0.50 were predominantly associated with healthy participants, while scores above 0.75 were strongly associated with PD. Intermediate scores represented a clinically uncertain zone in which overlap between diagnostic groups increased.

## 4. Discussion

This is a proof-of-concept study establishing clinical validity — that a reliable eye-tracking signal for PD exists and is detectable using a reproducible method. SaccadeDX™, analyzed with a two-level cascade classifier, detected PD versus non-PD participants with AUC = 0.866 and accuracy = 83.5%, meeting the prospectively specified accuracy threshold and the proof-of-concept AUC benchmark. The classifier provides a fully auditable scoring equation and conservative cross-validated performance estimates likely to replicate.

### 4.1 Biological Plausibility of Selected Features

The features that survived elastic-net selection map onto neuroanatomic circuits with established involvement in movement-disorder pathophysiology. Vertical saccade curvature (Level 1) reflects integrity of the rostral interstitial nucleus of the medial longitudinal fasciculus (riMLF) and superior colliculus, structures with documented degeneration in tauopathies and early targets of α-synuclein pathology in PD. Pursuit deficits (Level 2: fast and slow pursuit metrics) implicate cortico-pontine-cerebellar circuits, which are differentially affected in PD versus PSP and essential tremor [4,24,27]. Fixation main-sequence slope abnormalities (Level 2) reflect basal-ganglia–superior-colliculus interactions modulated by dopamine; the positive coefficient (PD > OM) is consistent with the PD-specific saccadic hypometria literature [21,25]. Right-OKN slow-phase abnormalities and increased vertical saccade misses (Level 2) reflect cerebellar and brainstem gaze-control disruption, distinguishing OM (which includes ataxic and cerebellar phenotypes) from PD. These mappings indicate that the discriminative signal tracks known neuropathology rather than statistical artifact. The full network of cortical, basal-ganglia, brainstem, and cerebellar nodes underlying these measurements is shown schematically in Supplementary Figure S1. Detailed feature-importance rankings and neural pathway annotations are provided in Supplementary Table S4.

### 4.2 Positioning Within the Eye-Tracking Literature in Parkinson’s Disease

Diotaiuti et al. recently reviewed the PD eye-tracking literature and concluded that eye movement abnormalities are increasingly recognized as early, objective biomarkers of PD, reflecting both motor and cognitive dysfunction [7]. Their synthesis organized the literature around the same domains represented in the present feature set: saccadic impairment, fixation instability, smooth-pursuit dysfunction, and pupillary/autonomic abnormalities [7]. The review further emphasized that these oculomotor markers may support early diagnosis, disease monitoring, cognitive assessment, and rehabilitation, but that broader clinical adoption requires standardized assessment protocols and validation of predictive models [7].

The present study fits directly into that translational gap. SaccadeDX™ does not rely on a single oculomotor sign or an unvalidated black-box score. Instead, it collects multiple eye-movement domains in a brief binocular protocol and applies a reproducible, two-level cascade classifier. Level 1 addresses the clinically broad question of whether a participant differs from healthy controls, whereas Level 2 addresses the harder differential-diagnostic question of whether the movement-disorder signal is more consistent with PD or with other movement disorders. This distinction is important because the review literature shows that many oculomotor abnormalities occur across neurodegenerative disorders, including PSP, Huntington’s disease, ataxia, and MSA, rather than being uniquely diagnostic of PD [7].

The selected features also align with the domains emphasized in the review. Level-1 discriminators include vertical saccade curvature, fixation instability measures, pupillometry, and gain-related metrics, consistent with the review’s description of hypometric saccades, delayed or dysregulated saccadic control, square-wave jerks, and autonomic changes as PD-relevant oculomotor markers [7]. Level-2 discriminators emphasize smooth-pursuit amplitude and catch-up saccades, fixation main-sequence slope, optokinetic slow-phase duration, and vertical saccade misses, all of which are biologically plausible for distinguishing idiopathic PD from PSP, cerebellar disorders, essential tremor, and other movement disorders represented in the OM group [1,5,7,11,14,16–18,24].

Thus, the added value of the current device is not that it identifies a previously unknown eye-movement abnormality. Its contribution is operational: it converts a diverse set of known oculomotor markers into a standardized, auditable, cross-validated classification framework. This is consistent with the review’s call for standardized clinical protocols and validated predictive models, while preserving clinical interpretability through explicit feature-level summaries rather than relying solely on opaque machine-learning output [7].

### 4.3 Comparison with Prior Exploratory Analyses

An earlier exploratory analysis of the same dataset, in which a large grid of model configurations was searched, produced a higher apparent Level-2 AUC. That value reflected optimism from configuration search and is not the basis for inference in this manuscript. The cross-validated estimate of 0.670 is the conservative, replication-oriented number, and we report it without modification. We anticipate that the planned multi-center confirmatory study will permit a fully Bayesian re-analysis with adequate posterior identification, providing posterior distributions and credible intervals for diagnostic performance and allowing uncertainty to be quantified directly rather than inferred from cross-validated point estimates.

### 4.4 Clinical Relevance

The utility of this testing lies in its ease of use and lack of reliance on clinical interpretation of findings. While testing eye movements in the clinic is relatively straightforward, there are many subtleties that require an in-depth understanding of the underlying neuroanatomy and disease processes; further, some findings may be difficult to quantify or detect by direct visualization. Regarding PD, there may be small square-wave jerks but not large macrosaccades [1,11,19]. Saccades in PD may be hypometric, with self-paced saccades more affected than reflexive saccades [21,25]. In PSP, eye movement changes are a defining feature of the disease [12]. Macro square-wave jerks, slowing of vertical saccades (particularly downward), and eventual supranuclear gaze palsy help to define this disorder clinically; however, only about one quarter of PSP patients will have sufficient eye movement changes at presentation to meet diagnostic criteria [1,5,11,16,17]. Automated testing of eye movements with the ability to detect small changes may be useful in early disease, particularly when faced with a participant presenting with parkinsonism.

Huntington’s disease patients also have eye movement abnormalities that start early in the disease course and progress with other motor impairment [1]. These changes include prolonged saccade latency (oculomotor apraxia), saccadic intrusion to new visual targets, and increased errors in anti-saccadic eye movements [14]. Patients with ataxia have dysmetric saccades and gaze-evoked nystagmus typical of cerebellar disorders [4]. Patients with essential tremor may have subtle eye movement changes suggestive of a cerebellar disorder and can be discriminated from tremor-dominant PD [24]. Distinguishing between these different types of movement disorder is beyond the scope of the current study due to low subject numbers in each subgroup; further study will help to clarify this aspect.

A practical advantage of the current framework is that the selected variables are clinically interpretable. The device may therefore be understood as a structured measurement system for known oculomotor abnormalities rather than as an unexplained statistical classifier. This is important for clinical adoption because the same review literature that supports eye tracking as a promising biomarker also highlights the need for standardization, model validation, and clinically meaningful interpretation before routine implementation [7].

### 4.5 Limitations

Single-center recruitment limits generalizability. The sample is sufficient to establish signal existence but not population-level calibration. Absence of medication-timing data is a meaningful gap that should be addressed in the confirmatory study. The OM group is heterogeneous and insufficient in number for between-disorder comparisons within OM. None of these constraints undermines the proof-of-concept conclusion: a reliable, reproducible eye-tracking signal for PD exists and can be detected by an objective, automated method.

Interpretation of individual selected features should be approached cautiously. Many oculomotor measurements are highly correlated and may represent overlapping physiological processes. In such settings, elastic-net regularization often selects one representative feature from a correlated cluster while shrinking related variables toward zero. Consequently, differences such as selection of quick-phase velocity in one direction and quick-phase magnitude in the opposite direction should not necessarily be interpreted as distinct biological mechanisms. Rather, the broader oculomotor domains represented by the selected features—including fixation stability, smooth pursuit, optokinetic responses, and saccadic control—are likely to be more stable than any individual variable. The planned multi-center confirmatory study will evaluate feature-selection stability through bootstrap resampling, repeated cross-validation, and external validation.

### 4.6 Future Directions

The complexity of the planned multi-center confirmatory study — with the cohorts stratified across age, symptom duration (which may not be coextensive with disease duration), sex, and duration of clinical signs to accommodate the heterogeneity of presentation within and across movement-disorder subtypes — is expected to require on the order of 300–500 patients. Sample-size determination will use the Hanley–McNeil approximation [10].

## 5. Conclusions

Clinical validity is established: a reproducible eye-tracking signal for Parkinson’s disease can be detected by an objective, automated method using a two-level cascade classifier (end-to-end AUC = 0.866, accuracy = 83.5%). The discriminative features map onto established oculomotor neuroanatomic circuits, supporting biological plausibility. A multi-center confirmatory study, stratified across age, symptom duration, sex, and duration of clinical signs, is the appropriate next step before assessment of clinical utility and broader diagnostic implementation.

## Data Availability

Data Availability Statement
De-identified participant-level data underlying this study are not publicly available due to participant privacy protections and contractual restrictions. Aggregate summary data, model coefficients, and analysis code are available from the corresponding author upon reasonable request and subject to institutional and sponsor approval.

https://clinicaltrials.gov/study/NCT04925622

## Author Contributions

J.M.M.: Methodology, Software, Formal analysis, Visualization, Writing – original draft. S.A.: Investigation, Data curation, Writing – review & editing. H.R.: Conceptualization, Methodology, Software. R.W.: Conceptualization, Project administration, Funding acquisition. H.A.S.: Investigation, Resources, Supervision, Writing – review & editing. All authors approved the final manuscript.

## Data Availability

De-identified participant-level data underlying this study cannot be shared publicly due to participant privacy protections and contractual restrictions. Aggregate summary data, the full coefficient table, and analysis code are available from the corresponding author upon reasonable request and subject to institutional and sponsor approvals.

## Declaration of generative AI and AI-assisted technologies in the manuscript preparation process

During the preparation of this work the authors used Claude (Anthropic) in order to assist with manuscript drafting, statistical code review, and reference reconciliation. After using this tool/service, the authors reviewed and edited the content as needed and take full responsibility for the content of the published article.

## Acknowledgements

None

## Supplementary Material

[Figure S5 about here]

[Table S1 about here]

[Table S2 about here]

[Table S3 about here]

[Table S4 about here

Figures 1 – 4**, S1**

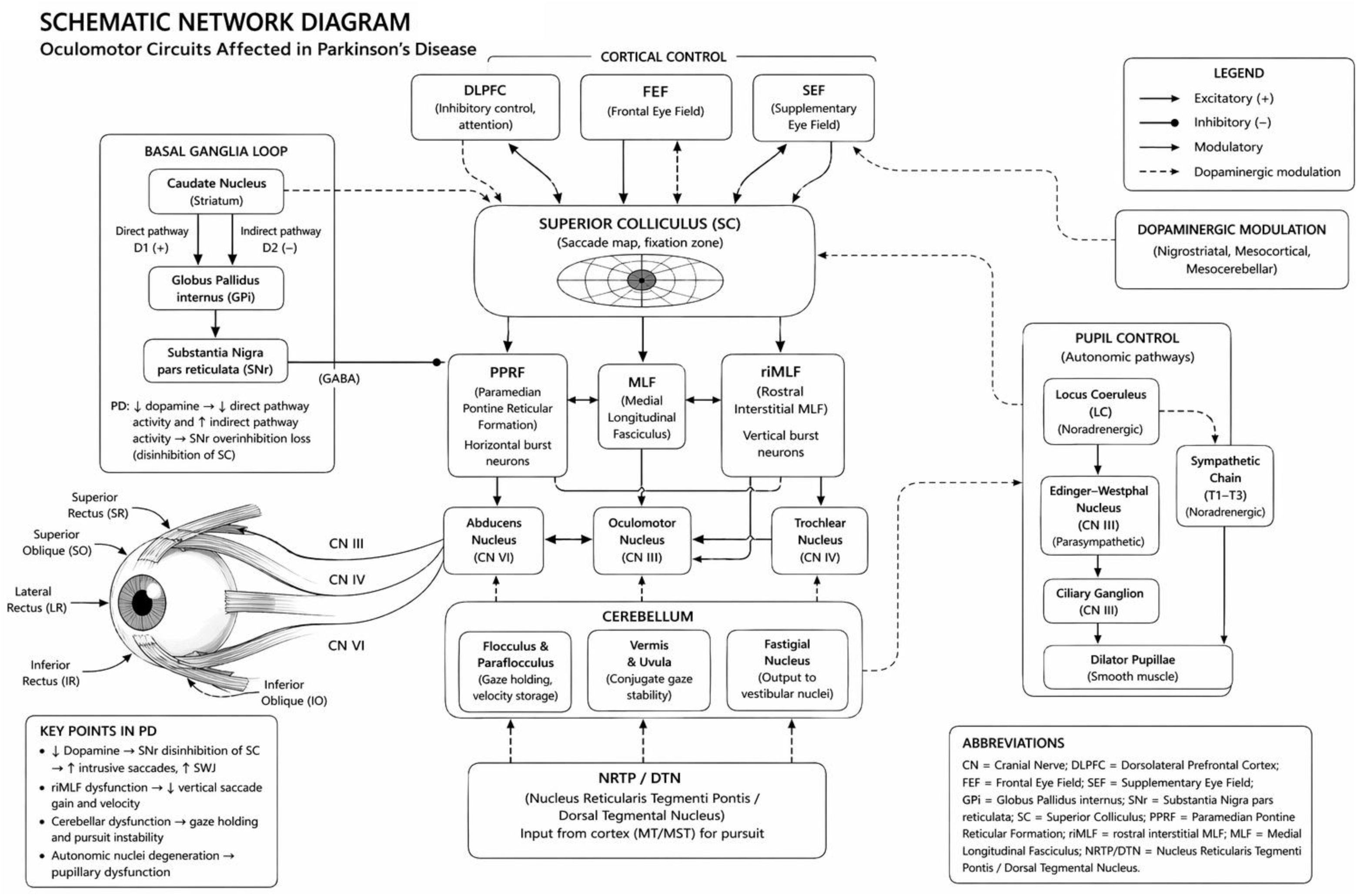

