## Supplemental Table 1. for "Automated Eye-Tracking for Parkinson’s Disease Diagnosis: A Proof-of-Concept Cascade Classifier Study Establishing Clinical Validity"

**Supplementary Table S1.** Level-1 discriminating eye-tracking features: healthy controls vs. movement disorder.

| Rank | Feature | Clinical meaning | Circuit / process | $\lvert\beta\rvert$ |
|---|---|---|---|---|
| 1 | Vertical saccade curvature | Abnormal vertical trajectory control | Caudate-SNr-SC; riMLF | 0.393 |
| 2 | Fixation harmonic velocity | Fixation instability / microsaccadic velocity | SNr-SC; cerebellum | 0.080 |
| 3 | Vertical saccade gain | Vertical hypometria | riMLF; SNr-SC | 0.041 |
| 4 | Vertical gain deficit | Engineered measure of abnormal gain | riMLF; basal ganglia-collicular | 0.038 |
| 5 | Pupillary baseline | Autonomic / arousal signal | LC; Edinger-Westphal | 0.018 |
| 6 | SWJ × OKN slow phase | Fixation plus optokinetic abnormality | SNr-SC plus NOT/DTN-cerebellar | 0.013 |
| 7 | Fixation saccade magnitude | Intrusive saccades during fixation | SNr-SC; FEF | 0.012 |
| 8 | Fixation saccade count | Frequency of fixation intrusions | SNr-SC fixation network | 0.010 |
| 9 | Pupil contraction delay | Delayed pupillary light reflex | Edinger-Westphal / parasympathetic | 0.008 |
| 10 | Fast-pursuit saccade magnitude | Catch-up saccades during pursuit | Putamen-cerebellar pursuit; SC | 0.007 |
