## Supplemental Table 2. for "Automated Eye-Tracking for Parkinson’s Disease Diagnosis: A Proof-of-Concept Cascade Classifier Study Establishing Clinical Validity"

**Supplementary Table S2.** Level-2 discriminating eye-tracking features: Parkinson's disease vs. other movement disorders.

| Rank | Feature | Clinical meaning | Circuit / process | $\lvert\beta\rvert$ |
|---|---|---|---|---|
| 1 | Vertical saccade misses † | Failed vertical targeting; OM > PD | riMLF; SNr-SC | 1.313 |
| 2 | Fast-pursuit saccade rate | Frequent catch-up saccades; OM > PD | Cerebellar-brainstem pursuit; SC | 0.681 |
| 3 | Slow-pursuit saccade rate | Corrective pursuit saccades; OM > PD | MT/MST-putamen-cerebellum | 0.561 |
| 4 | Left OKN quick-phase velocity | Optokinetic quick phase; PD > OM | PPRF; SC-PPRF | 0.434 |
| 5 | Right OKN slow-phase duration † | Velocity-storage / OKN signal; OM > PD | NOT/DTN; cerebellum | 0.315 |
| 6 | Fixation main-sequence slope † | Microsaccade velocity-amplitude relation; PD > OM | SNr-SC; caudate-SNr | 0.314 |
| 7 | Fast-pursuit right-eye amplitude † | Pursuit amplitude abnormality; OM > PD | MT/MST-putamen; corticobulbar/cerebellar | 0.265 |
| 8 | Slow-pursuit saccade magnitude † | Catch-up magnitude; OM > PD | MT/MST-putamen; flocculus | 0.247 |
| 9 | Left OKN slow-phase duration | Optokinetic slow-phase signal; OM > PD | NOT/DTN-cerebellar OKN | 0.181 |
| 10 | Fast-pursuit RMSE | Irregular pursuit; PD > OM | Putamen-cerebellar smoothing | 0.172 |

† 95% confidence interval excludes zero.
