## Supplemental Table 3. for "Automated Eye-Tracking for Parkinson’s Disease Diagnosis: A Proof-of-Concept Cascade Classifier Study Establishing Clinical Validity"

**Supplementary Table S3.** Full feature inventory with selection status across both cascade levels.

All 97 candidate features (75 raw oculomotor measurements plus 22 engineered features). Rows showing "Yes" in L1 or L2 selected columns indicate features retained by the elastic-net penalty.

| Feature | L1 selected | $\|\beta\|$ L1 | L2 selected | $\|\beta\|$ L2 |
|---|---|---|---|---|
| feat_eng_fix_instability | — | — | — | — |
| feat_eng_fix_latency | — | — | — | — |
| feat_eng_gain_deficit_ratio | — | — | — | — |
| feat_eng_horiz_gain_deficit | — | — | — | — |
| feat_eng_mainseq_ratio | — | — | — | — |
| feat_eng_okn_lateral | — | — | — | — |
| feat_eng_okn_qp_mean | — | — | — | — |
| feat_eng_okn_qs_ratio | — | — | — | — |
| feat_eng_okn_saccade_prod | — | — | — | — |
| feat_eng_okn_sp_mean | — | — | — | — |
| feat_eng_pupil_asymmetry | — | — | — | — |
| feat_eng_pupil_swj | — | — | — | — |
| feat_eng_pursuit_catchup_x_deficit | — | — | — | — |
| feat_eng_pursuit_lateral | — | — | — | — |
| feat_eng_pursuit_okn_ratio | — | — | — | — |
| feat_eng_pursuit_rmse_mean | — | — | — | — |
| feat_eng_swj_okn | Yes | 0.013 | — | — |
| feat_eng_swj_vert_product | — | — | — | — |
| feat_eng_swj_x_okn_sp | — | — | — | — |
| feat_eng_vert_gain_deficit | Yes | 0.038 | — | — |
| feat_eng_vert_horiz_ratio | — | — | — | — |
| feat_eng_vert_severity | — | — | — | — |
| fixation_bcea | — | — | — | — |
| fixation_harmonicvelocity | Yes | 0.080 | — | — |
| fixation_horizontal_std | — | — | — | — |
| fixation_horizvertcorr | — | — | — | — |
| fixation_mainsequenceslope | — | — | Yes | 0.314 |
| fixation_meanbinoculardisparity | — | — | — | — |
| fixation_meannonswjverticalcomponent | — | — | — | — |
| fixation_meanpeakvelocity | — | — | — | — |
| fixation_meansaccademagnitude | Yes | 0.012 | — | — |
| fixation_meanswjverticalcomponent | — | — | Yes | 0.135 |
| fixation_numberofsaccades | Yes | 0.010 | — | — |
| fixation_recordingduration | — | — | — | — |
| fixation_swjrate | — | — | — | — |
| fixation_vertical_std | — | — | — | — |
| fixation_verticalvarianceproportion | — | — | — | — |
| group | — | — | — | — |
| horizontal.saccades_averagegain | — | — | — | — |
| horizontal.saccades_averagelatency | — | — | — | — |
| horizontal.saccades_curvatureleft | — | — | — | — |
| horizontal.saccades_curvatureright | — | — | — | — |
| horizontal.saccades_mainsequenceslope | — | — | — | — |
| horizontal.saccades_numberofmisses | — | — | — | — |
| leftokn_quickphases_magnitude | — | — | — | — |
| leftokn_quickphases_peakvelocity | — | — | Yes | 0.434 |
| leftokn_slowphases_duration | — | — | Yes | 0.181 |
| leftokn_slowphases_magnitude | — | — | — | — |
| pupillometry_baseline | Yes | 0.018 | — | — |
| pupillometry_lefteye_baseline | — | — | — | — |
| pupillometry_lefteye_maxcontraction | — | — | — | — |
| pupillometry_lefteye_maxcontractiondelay | Yes | 0.008 | — | — |
| pupillometry_maxcontraction | — | — | — | — |
| pupillometry_maxcontractiondelay | — | — | — | — |

| Feature | L1 selected | $\lvert\beta\rvert$ L1 | L2 selected | $\lvert\beta\rvert$ L2 |
|---|---|---|---|---|
| pupillometry_righteye_baseline | — | — | — | — |
| pupillometry_righteye_maxcontraction | — | — | — | — |
| pupillometry_righteye_maxcontractiondelay | — | — | — | — |
| pursuitfast_amplitude | — | — | — | — |
| pursuitfast_delay | — | — | — | — |
| pursuitfast_frequency | — | — | — | — |
| pursuitfast_lefteye_amplitude | — | — | — | — |
| pursuitfast_lefteye_delay | — | — | — | — |
| pursuitfast_lefteye_frequency | — | — | — | — |
| pursuitfast_lefteye_rmse | — | — | Yes | 0.104 |
| pursuitfast_righteye_amplitude | — | — | Yes | 0.265 |
| pursuitfast_righteye_delay | — | — | — | — |
| pursuitfast_righteye_frequency | — | — | — | — |
| pursuitfast_righteye_rmse | — | — | — | — |
| pursuitfast_rmse | — | — | Yes | 0.172 |
| pursuitfast_saccademagnitude | Yes | 0.007 | — | — |
| pursuitfast_saccaderate | — | — | Yes | 0.681 |
| pursuitfast_velocitygain | — | — | — | — |
| pursuitslow_amplitude | — | — | — | — |
| pursuitslow_delay | — | — | — | — |
| pursuitslow_frequency | — | — | — | — |
| pursuitslow_lefteye_amplitude | — | — | Yes | 0.105 |
| pursuitslow_lefteye_delay | — | — | — | — |
| pursuitslow_lefteye_frequency | — | — | — | — |
| pursuitslow_lefteye_rmse | — | — | — | — |
| pursuitslow_righteye_amplitude | — | — | — | — |
| pursuitslow_righteye_delay | — | — | — | — |
| pursuitslow_righteye_frequency | — | — | — | — |
| pursuitslow_righteye_rmse | — | — | — | — |
| pursuitslow_rmse | — | — | — | — |
| pursuitslow_saccademagnitude | — | — | Yes | 0.247 |
| pursuitslow_saccaderate | — | — | Yes | 0.561 |
| pursuitslow_velocitygain | — | — | — | — |
| rightokn_quickphases_magnitude | — | — | Yes | 0.111 |
| rightokn_quickphases_peakvelocity | — | — | — | — |
| rightokn_slowphases_duration | — | — | Yes | 0.315 |
| rightokn_slowphases_magnitude | — | — | — | — |
| smooth.convergence_npc | — | — | — | — |
| vertical.saccades_averagegain | Yes | 0.041 | — | — |
| vertical.saccades_averagelatency | — | — | — | — |
| vertical.saccades_curvatureleft | Yes | 0.393 | — | — |
| vertical.saccades_curvatureright | — | — | — | — |
| vertical.saccades_mainsequenceslope | — | — | — | — |
| vertical.saccades_numberofmisses | — | — | Yes | 1.313 |

Total features: 97. Selected for Level 1: 10 of 97. Selected for Level 2: 14 of 97. "—" indicates the feature was shrunk to zero by the elastic-net penalty at $\lambda_{1se}$. $\lvert\beta\rvert$ values are shown for selected features.
