## Supplemental Table 4. for "Automated Eye-Tracking for Parkinson’s Disease Diagnosis: A Proof-of-Concept Cascade Classifier Study Establishing Clinical Validity"

**Supplementary Table S4.** Feature Importance and Neural Pathway Annotation — Two-Level Cascade Classifier.

Importance = |coefficient| from logistic regression with elastic-net feature selection. † = 95% confidence interval excludes zero (selected features only). Evidence rating reflects quality of pathway-to-measure linkage in published literature, not feature classification performance.

| | |
|---|---|
| ●●● | Established — multiple independent studies |
| ●●○ | Inferred — consistent with known pathway, limited direct evidence |
| ●○○ | Not established — tract-to-measure link not supported by sufficient evidence; annotation withheld |

*S4a. Level 1 — Healthy Controls vs Movement Disorders (top 20 of 97 features tested; 10 selected by elastic net).*

| Rank | Feature | $\|\beta\|$ | Direction | Mechanism | Evidence |
|---|---|---|---|---|---|
| 1 | Vertical Saccades Curvature Left | 0.393 | ↓ in MD | Striatal dopamine depletion disrupts the inhibitory gating of curved saccade trajectories; curvature reflects misdirected collicular burst | ●●● |
| 2 | Fixation Harmonic Velocity | 0.080 | ↑ in MD | Harmonic velocity components of fixation micro-saccades reflect SNr disinhibition plus cerebellar velocity-storage instability | ●●○ |
| 3 | Vertical Saccades Average Gain | 0.041 | ↓ in MD | Vertical hypometria reflects riMLF burst neuron impairment and/or reduced SC disinhibition via the SNr pathway; more pronounced vertically than horizontally in PD | ●●● |
| 4 | [Eng] Vertical Gain Deficit | 0.038 | ↑ in MD | Engineered: distance of vertical gain from perfect (1.0); amplifies the riMLF signal already captured by average gain | ●●○ |
| 5 | Pupillometry Baseline | 0.018 | ↑ in MD | *Tracts:* locus coeruleus, Edinger-Westphal nucleus, sympathetic chain (T1-T3). Autonomic dysregulation (Braak stages 2–4) causes mydriasis via LC noradrenergic and EWN parasympathetic degeneration | ●●○ |
| 6 | [Eng] SWJ × OKN Slow Phase Inverse | 0.013 | ↑ in MD | Combined SNr disinhibition (elevated SWJ) and accessory optic system impairment (reduced OKN slow phase); product term captures co-occurrence | ●●○ |
| 7 | Fixation Mean Saccade Magnitude | 0.012 | ↑ in MD | Elevated micro-saccade amplitude during fixation reflects reduced SNr inhibitory tone on SC; FEF top-down control also impaired | ●●○ |
| 8 | Fixation Number of Saccades | 0.010 | ↑ in MD | Intrusive saccades during fixation; elevated count indicates SNr disinhibition allowing SC burst neuron activation without voluntary intent | ●●● |
| 9 | Pupillometry Left Eye Max Contraction Delay | 0.008 | ↑ in MD | *Tracts:* Edinger-Westphal nucleus (parasympathetic), Braak stage 3–4. EWN degeneration slows pupillary light reflex contraction; asymmetric Lewy body burden may produce left–right differences | ●●○ |
| 10 | Pursuit Fast Saccade Magnitude | 0.007 | ↓ in MD | Catch-up saccade magnitude during fast pursuit reflects D2-pathway putaminal involvement reducing pursuit gain | ●●○ |
| 11–20 | Remaining 10 features (imp 0.004–0.006) | 0.004–0.006 | Various | Coefficients shrunken to near zero by elastic-net penalty ($\|\beta\| < 0.007$). Below selection threshold. Pathway annotation not warranted. | ●○○ |

*S4b. Level 2 — Parkinson's Disease vs Other Movement Disorders (top 20 of 97 features tested; 14 selected by elastic net).*

| Rank | Feature | $\|\beta\|$ | Direction | Mechanism | Evidence |
|---|---|---|---|---|---|
| 1 | Vertical Saccades Number of Misses † | 1.313 | OM > PD | Complete failure of vertical saccades (misses) indicates riMLF destruction — characteristic of PSP in the OM group. Idiopathic PD shows hypometria but not complete failure, making this a PSP/CBD vs PD discriminator | ●●● |
| 2 | Pursuit Fast Saccade Rate | 0.681 | OM > PD | *Tracts:* SC disinhibition; cerebellar-brainstem (ataxia). High catch-up saccade rate during fast pursuit in OM reflects cerebellar loop failure (ataxia subgroup) and/or SC hyperactivation — both more severe in cerebellar/ataxic OM than idiopathic PD | ●●○ |
| 3 | Pursuit Slow Saccade Rate | 0.561 | OM > PD | Same pathway logic as fast; slow pursuit saccade rate elevated in PSP/CBD and ataxia more than idiopathic PD due to combined corticobulbar and cerebellar pathway disruption | ●●○ |
| 4 | Left OKN Quick Phases Peak Velocity | 0.434 | PD > OM | OKN quick phase velocity mirrors saccade velocity. In idiopathic PD, PPRF is relatively preserved early; in PSP and ataxic OM, PPRF degeneration is more severe — explaining PD > OM direction | ●●○ |
| 5 | Right OKN Slow Phases Duration † | 0.315 | OM > PD | OKN slow phase duration reflects velocity storage time constant. Cerebellar OM conditions (ataxia, PSP cerebellar variant) impair floccular velocity storage more severely than idiopathic PD | ●●● |
| 6 | Fixation Main Sequence Slope † | 0.314 | PD > OM | The main sequence (velocity/amplitude ratio) slope during fixation micro-saccades is elevated in PD specifically due to SNr disinhibition; essential tremor in OM group has normal fixation main sequence | ●●● |
| 7 | Pursuit Fast Right Eye Amplitude † | 0.265 | OM > PD | Pursuit amplitude reduction in OM reflects more extensive corticobulbar disruption in PSP/CBD than in idiopathic PD at equivalent disease stage. CBD additionally impairs the cortical motor planning of smooth pursuit | ●●● |
| 8 | Pursuit Slow Saccade Magnitude † | 0.247 | OM > PD | Catch-up saccade magnitude during slow pursuit elevated in ataxic OM (cerebellar pathway failure) and PSP (brainstem); smaller in idiopathic PD where putaminal D2 pathway is primary lesion | ●●○ |
| 9 | Left OKN Slow Phases Duration | 0.181 | OM > PD | Same velocity-storage mechanism as Right OKN slow phase duration (rank 5). Bilateral OKN slow phase elevation in cerebellar OM; unilateral asymmetry possible in early PD (lateralised dopamine loss) | ●●● |
| 10 | Pursuit Fast RMSE | 0.172 | PD > OM | *Tracts:* cerebellum (flocculus/vermis) — smoothing failure; putamen (D2). Higher RMSE in PD reflects putaminal D2 pathway disruption of pursuit smoothness; paradoxically lower in some OM conditions (PSP/CBD) where pursuit is simply absent rather than jerky | ●●○ |
| 11–20 | Remaining 10 features (imp 0.071–0.135) | 0.071–0.135 | Various | Features 11–20 have meaningful importances (0.071–0.135) but wider confidence intervals. Pathway linkage for individual features in this range is not established with sufficient confidence to assert in a peer-reviewed table. | ●○○ |

Abbreviations: CBD, corticobasal degeneration; DTN, dorsal tegmental nucleus; FEF, frontal eye fields; LC, locus coeruleus; MLF, medial longitudinal fasciculus; MT/MST, middle temporal / medial superior temporal cortex; NOT, nucleus of the optic tract; PPRF, paramedian pontine reticular formation; PSP, progressive supranuclear palsy; riMLF, rostral interstitial nucleus of the MLF; SC, superior colliculus; SNr, substantia nigra pars reticulata; SWJ, square-wave jerk. Tract evidence based on established oculomotor anatomy and PD/parkinsonism literature; ratings reflect strength of direct empirical linkage to the specific measure, not general pathway involvement.
